# A systematic review and meta analysis of the valve of SHOX2 and RASSF1A gene methylation in bronchial elveslar lavage fluid in the diagnosis of lung cancer

**DOI:** 10.1101/2022.05.31.22275806

**Authors:** Shanyang Su, Yanling Huang, Xiang Lu, Wenjia Li, Yongshun Li, Jihong Zhou

**Author notes:** Correspondence: Jihong Zhou, Dr., Shenzhen Bao’an Traditional Chinese Medicine Hospital,Guangzhou University of Chinese Medicine, 518101, Guangdong, China. Data sharing not applicable to this article as no datasets were generated or analyzed during the current study. This research didn’t receive grants from any funding agency in the public, commercial or not-for-profit sectors.

## Abstract

**Introduction:** The incidence of lung cancer worldwide has been increasing in recent years, and the latest cancer data published by the WHO International Agency for Research on Cancer in 2021 shows that lung cancer remains the cancer with the highest mortality rate. The sensitivity of early screening methods for lung cancer is not ideal. Because early lung cancer is mostly located in nodules with a diameter of 1 cm, it is difficult to obtain a definite pathological diagnosis from living tissue specimens. Therefore, it is necessary to find less invasive and effective tests to help detect lung cancer early. Therefore, the purpose of this systematic review (SR) and meta-analysis will be to analyze and explore the correlation between promoter methylation of SHOX2 and RASSF1A genes and lung cancer, and to provide a reference for early clinical diagnosis.

**Methods and Analysis:** The relevant literature will be comprehensively searched in 4 international electronic databases (PubMed, Cochrane Library, EMBASE and Web of Science) and 4 Chinese electronic databases (CNKI, VIP, Wanfang, Chinese Biomedicine). We only included studies from inception until publication in May 2022. The primary outcome measure was the methylation rate of the SHOX2 and RASSF1A gene promoters in lung cancer tissue and normal lung tissue in the control group in lung cancer patients. Secondary outcome measures included methylation rates of promoters of SHOX2 and RASSF1A genes in different tissue samples. Two reviewers will conduct independent research selection, data extraction, data synthesis and quality assessment. The assessment of bias risk and data synthesis will be conducted using Review Manager 5.3 software. The Cochrane Collaboration’s Bias Risk Assessment Tool (QUADAS) will be used to assess the quality of the individual studies included.

**Discussion:** This systematic review will help to clarify the correlation between PROMOTER methylation of the SHOX2 and RASSF1A genes with lung cancer, providing clinical evidence for early clinical diagnosis.

**Trial registration number:** CRD42022330609 (PROSPERO)

## 1. Introduction

Primary bronchial lung cancer, referred to as lung cancer, is a malignant tumor that originates from the bronchial mucosa or glands. Lung cancer is insidious, mostly asymptomatic in the early stage, and clinical manifestations such as cough, sputum blood or hemoptysis, shortness of breath, fever, weight loss, chest pain, dyspnea, dysphagia, and hoarseness in the middle and late stages. Due to poor prognosis due to inadequate early diagnosis, studies have shown a five-year survival rate of only 19.8% in lung cancer patients^.[1]^ The latest cancer data published by WHO’s International Agency for Research on Cancer in 2021 shows that lung cancer remains the cancer with the highest mortality rate.^[2]^ In the early diagnosis and early intervention, two-thirds of patients can survive for at least 5 years, and some patients may benefit from long-term survival or even cure lung cancer.^[3, 4]^

Early diagnosis and early treatment of lung cancer is key to improving the five-year survival rate of lung cancer patients. The early screening methods of lung cancer are mainly tumor markers, cytology, imaging and tracheoscopy, but the sensitivity is not ideal, and it is very difficult to distinguish the benign and malignant lesions of small lung nodules. At present, the gold standard for clinical detection of lung cancer is pathological examination, because early lung cancer is mostly located in nodules with a diameter of 1 cm, it is difficult to obtain living tissue specimens to confirm the pathological diagnosis. There have been reports of cytological detection rates of bronchoalveolar lavage fluid in patients with lung cancer,^[5]^ second only to puncture biopsy and malignant pleural effusion cytopathological tests for the diagnosis of lung cancer.^[6]^ The combined detection of early lung cancer by shox2 and RASSF1A gene methylation is more effective than cytological testing, especially if the cytology test is unclear or negative, and can be used as a supplement to cytology tests.^[7]^ It can also improve the detection rate of tumors in small biopsy samples in patients with non-small cell lung cancer, especially when the pathological morphology of small biopsy is difficult to give a definitive diagnosis.^[8]^ The combined detection of SHOX2 and RASSF1A gene methylation in alveolar lavage fluid is significantly superior to single-index detection and cytologic detection of alveolar lavage fluid.^[9]^ Some patients with methylated positive lung nodules have been shown to present clinical indications for lung cancer after six months of follow-up, even though pathology and imaging do not support the diagnosis of lung cancer at this stage.^[10]^ The results show that the methylation of tumor suppressor genes such as SHOX2 and RASSF1A is closely related to the early diagnosis of lung cancer,^[11]^ and this study takes the tumor suppressor genes RASSF1A and SHOX2 as examples to comprehensively collect clinical studies between RASSF1A, SHOX2 gene methylation and lung cancer, and explore the correlation between rassF1A and SHOX2 gene promoter methylation and lung cancer, which provides a reference for early clinical diagnosis.

## 2. Materials and methods

### 2.1 Study registration

The study protocol has been registered in PROSPERO (registration number: CRD42022330609). Evaluation reports will be conducted in accordance with the preferred reporting items in the systematic review and meta-analysis guidelines.

### 2.2 Study design

#### 2.2.1 Type of participants

Lung cancer patients, patients with benign lung tumors (with a clear pathological diagnosis), and normal people will be included. There are no restrictions on gender, race, curriculum, ethnicity or country. Eligibility criteria: complete bronchoalveolar lavage fluid SHOX2, RASSF1A test with test results, with clear tissue and /or cell pathological results.)

#### 2.2.2 Type of interventions

Because this is a diagnostic meta, there are no interventions.

#### 2.2.3 Types of outcome measures

The primary outcome will include methylation rates of SHOX2 and RASSF1A gene promoters in lung cancer tissues of patients with lung cancer and normal lung tissues of control group.Secondary outcome will include methylation rates of SHOX2 and RASSF1A gene promoters in the different tissue samples.

#### 2.2.4 Inclusion and exclusion criteri

Studies that met all of the following requirements will be included:

1. Diagnosis: Lung cancer patients diagnosed with pathological diagnosis are the case group, and non-lung cancer patients (benign diseases or healthy people) are the control group;
2. Diagnostic method: The test to be evaluated is bronchoalveolar lavage fluid SHOX2, RASSF1A detection to diagnose lung cancer, with pathological diagnosis as the gold standard;
3. Participants were diagnosed with pulmonary nodules by clinicians based on diagnostic criteria in the original study;

At the same time, for multiple reports of the same study, we only include the most recent reports.

Studies that met one of the following requirements will be excluded:

1. Incomplete diagnostic data cannot be obtained;
2. Repeated publication and repeated detection;
3. Incomplete data cannot be extracted;
4. Case reports, animal experiments, qualitative studies, reviews or review articles.

### 2.3 Literature search strategy

We will search articles in four international electronic databases (PubMed, Cochrane Library, EMBASE, and Web of Science) and 4 Chinese electronic databases (China National Knowledge Infrastructure, VIP, Wanfang, and China Biology Medicine).All the publications until 30 April 2022 will be searched without any restriction of countries or article type. These studies must be published in English or Chinese. Reference list of all selected articles will independently screened to identify additional studies left out in the initial search. The literature search will be structured around search terms such as “non-small cell lung cancer, SHOX2 protein, human, RASSF1 protein, human, Bronchoalveolar Lavage Fluids” and adjust each database as needed. PubMed’s detailed search strategy is shown in Table 1. The search policy will also be used for any other electronic databases.

**Table 1.** Search strategy in PubMed.

| Serial number | Search items |
| --- | --- |
| #1 | Non Small Cell Lung Carcinoma[Mesh] |
| #2 | Carcinoma, Non Small Cell Lung[All Fields] |
| #3 | Carcinomas, Non-Small-Cell Lung[All Fields] |
| #4 | Lung Carcinoma, Non-Small-Cell[All Fields] |
| #5 | Lung Carcinomas, Non-Small-Cell[All Fields] |
| #6 | Non-Small-Cell Lung Carcinomas[All Fields] |
| #7 | Non-Small-Cell Lung Carcinoma[All Fields] |
| #8 | Nonsmall Cell Lung Cancer[All Fields] |
| #9 | Carcinoma, Non-Small Cell Lung[All Fields] |
| #10 | Non-Small Cell Lung Carcinoma[All Fields] |
| #11 | Non-Small Cell Lung Cancer[All Fields] |
| #12 | #1 or #2 - #11 |
| #13 | Bronchoalveolar Lavage Fluids[Mesh] |
| #14 | Lavage Fluids, Bronchial[All Fields] |
| #15 | Lavage Fluid, Bronchoalveolar[All Fields] |
| #16 | Lavage Fluids, Bronchoalveolar[All Fields] |
| #17 | Bronchial Alveolar Lavage Fluid[All Fields] |
| #18 | Pulmonary Lavage Fluid[All Fields] |
| #19 | Lavage Fluid, Pulmonary[All Fields] |
| #20 | Lavage Fluids, Pulmonary[All Fields] |
| #21 | Pulmonary Lavage Fluids[All Fields] |
| #22 | Lavage Fluid, Bronchial[All Fields] |
| #23 | Lavage Fluid, Lung[All Fields] |
| #24 | Lung Lavage Fluid[All Fields] |
| #25 | Lavage Fluids, Lung[All Fields] |
| #26 | Lung Lavage Fluids[All Fields] |
| #27 | Alveolar Lavage Fluid[All Fields] |
| #28 | Alveolar Lavage Fluids[All Fields] |
| #29 | Lavage Fluid, Alveolar[All Fields] |
| #30 | Lavage Fluids, Alveolar[All Fields] |
| #31 | Bronchial Lavage Fluid[All Fields] |
| #32 | Bronchial Lavage Fluids[All Fields] |
| #33 | #13 or #14 - #32 |
| #34 | SHOX2 protein, human[Mesh] |
| #35 | short stature homeobox 2 protein, human[All Fields] |
| #36 | OG12 protein, human[All Fields] |
| #37 | OG12X protein, human[All Fields] |
| #38 | #34 or #35 - #37 |
| #39 | RASSF1 protein, human[Mesh] |
| #40 | Ras association (RalGDS-AF-6) domain family 1 protein, human[All Fields] |
| #41 | RASSF1B protein, human[All Fields] |
| #42 | RASSF1C protein, human[All Fields] |
| #43 | #39 or #40 - #42 |
| #44 | #12 and #33 and #38 and #43 |

### 2.4 Study selection

In the first step of the data processing process, the retrieval strategy retrieves the titles and abstracts of all studies that are filtered for relevance, and the titles and abstracts of all studies that are clearly irrelevant will be discarded. If the results are not obviously irrelevant, the full text will be downloaded. In the second step, two members of the evaluation panel (Shanyang Su, Yanling Huang) will independently assess the eligibility of the study using the redefined inclusion and exclusion criteria. In addition, for studies that meet the inclusion criteria, reviewers will read the entire article to ensure that the entire study meets the criteria and are prepared to extract relevant information. Any disagreements on whether to include a particular study will be resolved through discussions between reviewers. (with a third author if necessary) to identify and resolve. For data on the missing part, we will contact the study authors for the missing data. The flowchart of all study selection procedures is shown in Figure 1.

**Figure 1:**
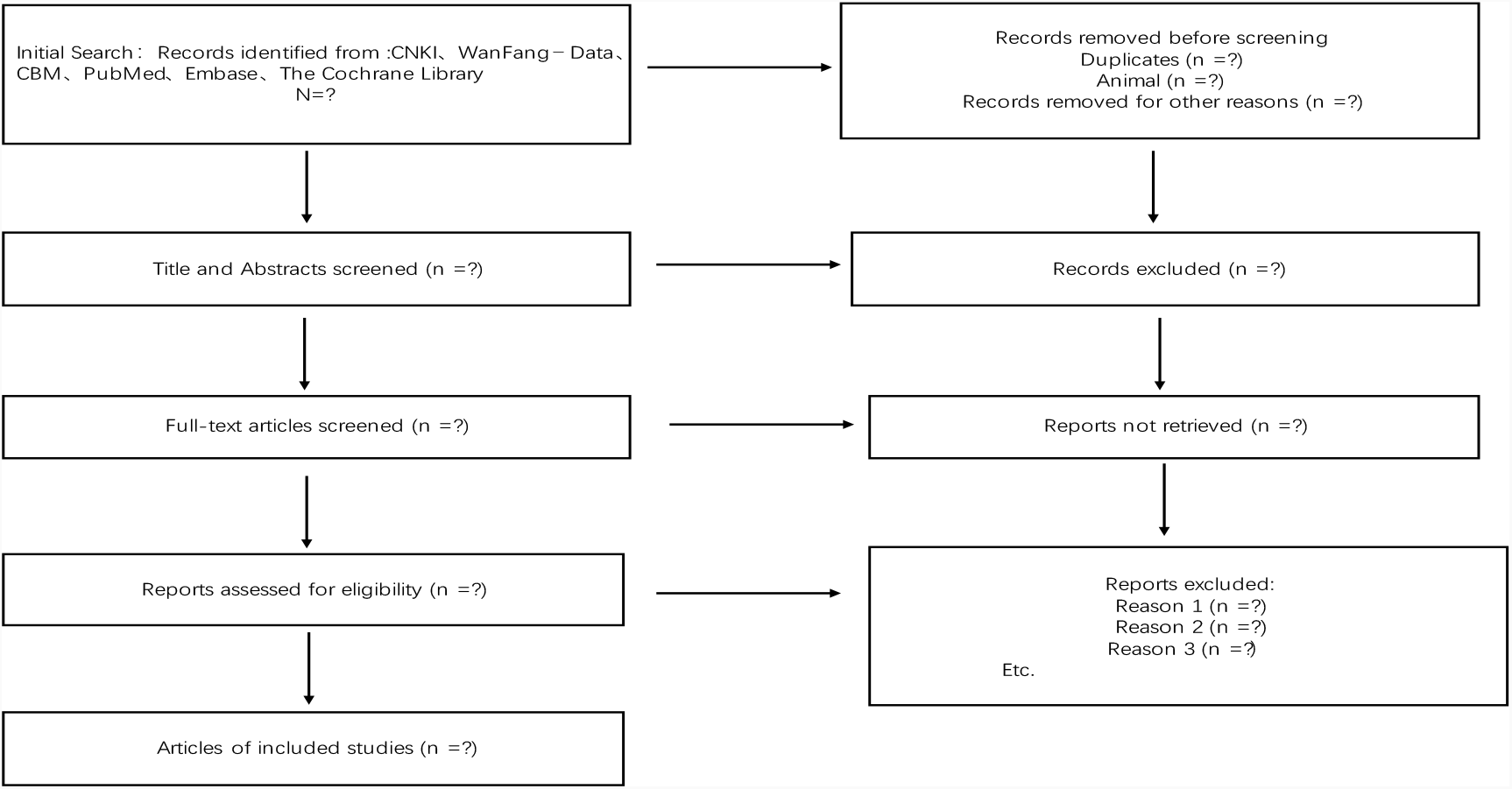
Flowchart of study selection procedures.

### 2.5 Data extraction

The information extracted by the 2 review team members from the relevant literature will include the first author name, study location, title, journal name, publication year, study environment, study population and participant demographics and baseline characteristics, detection methods, sample size included, methylation rate of promoters of shox2 and RASSF1A genes, number of SHOX2 positive cases, number of RASSF1A positive cases, number of SHOX2 and RASSF1A double positive cases, etc. used to assess the risk of bias. Two reviewers will extract the data independently; Discrepancies will be identified and resolved through discussion. If important data are missing, we will ask the study authors to provide the number of missing data.

### 2.6 Risk of bias Assessment

The bias risk assessment will involve 2 evaluators in the quality assessment process and any significant differences will be resolved through discussions to determine the final set of studies to be included. The two reviewers independently assessed the risk of bias in the included studies by taking into account the following characteristics: participant representation, gold standard reasonableness, intervals between trials, gold standard blind assessment, uncertain results and other sources of bias. In addition, the Cochrane Collaboration’s Bias Risk Assessment Tool (QUADAS) will be used to assess the quality of individual studies included.

### 2.7 Data synthesis

We will use Review Manager 5.3 software to carry out the quantitative synthesis if the included studies are sufficiently homogeneous. Mean difference or standardized means difference will be used for continuous data. Odds ratio (OR) will be used for the analysis of dichotomous data. Both we will give a 95% confidence interval (CI). In the case of homogeneous data, if I^2^≤50%, the fixed-effect model will be adopted for the meta-analysis. Otherwise, the sources of heterogeneity will be further analyzed. After excluding marked clinical heterogeneity, a random-effect model will be adopted to perform the meta-analysis. Sensitivity and bias risk analyses will also be performed.

#### 2.7.1. Analysis of subgroups

here are some planned subgroup analyses will be performed: different tissue samples of lung cancer (eg: tissue, BLAF, serum and pleural effusion), different pathological types of lung cancer (eg: squamous cell carcinoma and adenocarcinoma)

#### 2.7.2. Sensitivity analysis

Sensitivity analysis will be performed to determine the stability of the aggregated results by reducing 1 document at a time.

#### 2.7.3. Reporting bias analysis

Apply Stata 14.0 software to draw Deek’s funnel diagrams to identify publication bias in literature. Test level α=0.05. The rate parameter P<0.05, is considered to be publication biased

### 2.8. Quality of evidence

The Grading of Recommendations Assessment, Development, and Evaluation (GRADE) system will be used to assess the quality of the individual studies included. The five items (limitations, inconsistency, indirectness, imprecision, and publication bias) will be used to assess each outcome.

### 2.9. Patient and Public Involvement

Patients or the public were not involved in the design, or conduct, or reporting, or dissemination plans of our research

## 3. Ethics and dissemination

Because this systematic review will be conducted based on published research, there is no ethical approval requirement. The findings of this systematic review will be published in a peerreviewed journal.

## 4. Discussion

Abnormal methylation of the DNA gene promoter can be detected in a variety of malignancies and is expected to be a marker for early diagnosis. The occurrence of lung cancer is a complex process of multi-gene participation and multi-stage gradual evolution. The activation of oncogenes and the inactivation of tumor suppressors are closely related to the occurrence and development of lung cancer. Studies have found that epigenetics play an important role in the development of tumors. Gene methylation is a common epigenetic mechanism involved in regulating the occurrence, progression, and metastasis of lung cancer.^[12, 13]^ DNA methylation is currently the most well-studied form of modification in epigenetics. Normal gene methylation is necessary to maintain cell growth and metabolism, while abnormal DNA methylation can trigger diseases such as tumors. There are many changes in the degree of methylation of many unique tumor-related genes (including oncogenes and tumor suppressor genes) in the early stage of lung cancer, so the detection of DNA methylation is of great significance for the early diagnosis of cancer. SHOX2 is a member of the homologous box gene family that regulates gene expression and controller geneogenesis and cell differentiation. RASSF1A is an important tumor suppressor gene.^[14]^ RASSF1A is closely linked to the occurrence and development of a variety of malignant tumors, is a novel tumor suppressor gene, and RASSF1A is one of the most methylated and common genes in tumors.^[15]^ The in-depth exploration of SHOX2 and RASSF1A genes and their methylation will help promote basic and clinical research on tumors, and provide new directions and ideas for clinical diagnosis and treatment.

## Data Availability

Data sharing not applicable to this article as no datasets were generated or analyzed during the current study.

## Abbreviations

NSCLC: Non-Small Cell Lung Cancer
CI: confidence interval
PRISMA-P: Preferred Reporting Item for Systematic Review and Meta-analysis
OR: Odds ratio
TCM: traditional Chinese medicine
SHOX2: short stature home-obox 2
RASSF1A: ras associ-ation domain family 1

## 5. Author contributions

**Conceptualization:** Shanyang Su,Yanling Huang,Xiang Lu, Wenjia Li, Yongshun Li,Jihong Zhou.

**Data curation:** Xiang Lu, Jihong Zhou.

**Formal analysis:** Shanyang Su,Yanling Huang.

**Methodology:** Shanyang Su,Yanling Huang,Jihong Zhou..

**Project administration:**. Yongshun Li,Jihong Zhou.

**Resources:** Shanyang Su, Jihong Zhou.

**Software:** Shanyang Su, Yanling Huang.

**Visualization:** Xiang Lu,Wenjia Li.

**Writing – original draft:** Shanyang Su, Yanling Huang, Xiang Lu

**Writing – review & editing:** Yongshun Li, Jihong Zhou, Xiang Lu

---

Table 1: PubMed’s detailed search strategy

Figure 1: The flowchart of all study selection procedures

